# Exome sequencing in individuals with oculoauriculofrontonasal syndrome provides further evidence of a non-traditional inheritance model

**DOI:** 10.64898/2026.11.21.26368508

**Authors:** Henrique Regonaschi Serigatto, Nancy Mizue Kokitsu-Nakata, Siulan Vendramini-Pittoli, Cristiano Tonello, Maria Rita Passos-Bueno, Roseli Maria Zechi-Ceide

## Abstract

The oculoauriculofrontonasal syndrome is a rare condition characterized by ocular hypertelorism, broad nose, mandibular hypoplasia, preauricular tags, and ear anomalies. Despite presenting a clinically recognizable pattern defined by the overlap of craniofacial microsomia and frontonasal dysplasia, its etiology remains unknown. Males and females are equally affected, and there is no recurrence reported. Genetic investigations performed to date could not clarify this issue yet. To further investigate Mendelian inheritance and test the hypothesis of a possible monogenic cause, we performed exome sequencing in a case series of Brazilian individuals from a single center with the clinical diagnosis of oculoauriculofrontonasal syndrome, including both solo and trio-based exome sequencing. The results provided here further support a non-traditional inheritance model, requiring additional studies including multi-omics approaches to elucidate its etiology.

## Introduction

The oculoauriculofrontonasal syndrome (OAFNS; OMIM 601452) is a rare condition characterized by multiple congenital anomalies that arise from abnormal development of the frontonasal process and first and second pharyngeal arches (1,2). Clinically, it is recognizable by the overlapping features of frontonasal dysplasia (FND) and craniofacial microsomia (CFM). Main clinical findings include craniofacial asymmetry, ocular hypertelorism, epibulbar dermoid, broad nose, mandibular hypoplasia, ear anomalies, and preauricular tags (2). Almost a hundred cases have been reported to date; all cases are sporadic, and there is no sex predilection (2). Despite presenting a clinically recognizable pattern, its etiology is still unknown. Discordant twins have been reported, but the zygosity was unknown in all instances (2–4). Genetic investigations performed in a few individuals, including karyotyping and next-generation sequencing (NGS), have been unsuccessful so far (1,2,5–7). Therefore, in the face of previous negative results, a complex and non-traditional inheritance model, including postzygotic mosaicism, oligogenism, and nongenetic causes, has been suggested to explain the cause of OAFNS (1,2). Nonetheless, the absence of causal variants could be explained by the paucity of individuals analyzed. Thus, we aimed to perform exome sequencing (ES) in individuals with OAFNS to provide additional data and further support the current hypothesis regarding its etiology.

## Subjects and Methods

### Editorial Policies and Ethical Considerations

This study was approved by the Human Research Ethics Committee of HRAC/USP (protocol number 6.155.797; CAAE: 70420223.1.0000.5441) and performed in accordance with the Declaration of Helsinki. Written informed consent was obtained from each participant and/or parents before inclusion in this study. Permission to use images was also obtained.

### Sample characterization

This is a prospective study carried out at the Hospital for Rehabilitation of Craniofacial Anomalies, University of São Paulo (HRAC/USP), Bauru, São Paulo, Brazil. We included individuals with the clinical diagnosis of OAFNS, of both sexes and without age restriction. The diagnosis of OAFNS was made by clinical geneticists, based on the criteria proposed by Lehalle et al. (1)

### DNA extraction and exome sequencing

The DNA from each participant was extracted from peripheral blood samples (4 mL) using the FlexiGene DNA Kit (Qiagen, Hilden, Germany), according to the manufacturer’s protocol, and the DNA concentration was measured with the NanoDrop 2000 (Thermo Scientific, Wilmington, Delaware, USA) spectrophotometer. The DNA was then shipped to the facilities of the Human Genome and Stem Cell Research Center, Institute of Biosciences, University of São Paulo (IB/USP), where the ES was performed. ES library was prepared using the Nextera DNA Flex Library (Illumina, San Diego, California, USA) and captured with the xGen Exome Hyb Panel v2 (IDT). Cluster generation and sequencing were performed using NovaSeq 6000 v1.0 reagents (Illumina) on a NovaSeq 6000 System at 2×150bp. Sequencing data were processed with Picard Toolkit (http://broadinstitute.github.io/picard) and Burrows-Wheeler Aligner (BWA; http://bio-bwa.sourceforge.net/) to align sequences to the human genome reference GRCh38 and to perform quality filtering. Genome Analysis Toolkit (GATK; http://www.broadinstitute.org/gatk/) was used to perform variant calling, and variant annotation was done using ANNOVAR (https://annovar.openbioinformatics.org/), Ensembl Variant Effect Predictor (VEP)(8), and an in-house script for annotation of genomic variants in coding and noncoding regions. ES mean coverage was 149x, with 98% of bases presenting minimum coverage of 20x.

### Exome sequencing data analysis

The variant call files (VCF) of each case were individually uploaded to the online platform Franklin by Qiagen (https://franklin.genoox.com/). For the SNVs and small indels analysis, the variants were evaluated according to the ClinVar (https://www.ncbi.nlm.nih.gov/clinvar/) and the Leiden Open Variation Database (LOVD; https://www.lovd.nl/), populational frequency observed in the Genome Aggregation Database (gnomAD; https://gnomad.broadinstitute.org/) and the Online Archive of Brazilian Mutations (ABraOM; https://abraom.ib.usp.br/), in silico predictors and meta-predictors (REVEL, AlphaMissense, CADD, SIFT, FATHMM, MutationTaster, Polyphen-2, DANN, BayesDel, and SpliceAI) using thresholds defined by Pejaver et al.(9), and Online Mendelian Inheritance in Man (OMIM; https://www.omim.org/) associated phenotypes. Variants without any associated phenotypes were analyzed by prioritizing truncating variants, missense variants with high pathogenicity prediction scores, and available animal models. All variants were classified according to the ACMG/AMP guidelines (10) and its updates.

The following sets of filters were applied on Franklin: (i) Variants reported as pathogenic or likely pathogenic in ClinVar; (ii) Loss-of-function (LoF) variants including stop gain, stop loss, start gain, start loss, and frameshift variants, with minor allele frequency (MAF) < 0.01 and associated OMIM phenotype; (iii) Homozygous variants, with MAF < 0.01 and VAF (allele balance) > 0.8, associated or not with a OMIM phenotype; (iv) Variants filtered according to Human Phenotype Ontology (HPO; https://hpo.jax.org/) terms associated with each individual’s phenotype, with MAF < 0.01 and reported or not in ClinVar; (v) Variants with MAF < 0.01, medium quality by depth (10–20), VAF (20- 100%), with associated OMIM phenotype; (vi) Variants prioritized by Franklin’s AI, automatically dragged into the “workbench station”; (vii) Secondary findings according to the ACMG/AMP guidelines v3.3 (11).

Subsequently, a new search strategy was applied to further investigate causal variants: (i) MAF < 0.01; (ii) variants observed in exons, canonical splice sites, and splicing regions; (iii) including SNPs and indels; (iv) missense, frameshift, start loss, start gain, stop loss and stop gain variants; (v) variant within confidence intervals ranging from low to high; (vi) heterozygous and homozygous variants; (vii) missense variants with a REVEL score ≥ 0.644 or splice site variants with an aggregated prediction of a deleterious effect. In addition, data were manually analyzed by converting the VCF into sheets with Microsoft Excel software using the same strategy but replacing the meta-predictor REVEL with CADD (Combined Annotation-Dependent Depletion)(12), considering a threshold of ≥ 20 Phred score. Variants of interest were confirmed by visualization in the Integrative Genomics Viewer (IGV) software.

The copy number variants (CNVs) and structural variants (SVs) were also analyzed using Franklin, prioritizing variants with high-medium levels of confidence, using available data on DECIPHER (https://www.deciphergenomics.org/) and ClinVar, in addition to a comparison between the variants present in our cases with those cases from the literature presenting overlapping chromosomal regions and by analyzing the genes encompassed by those CNVs. CNVs and SVs with low confidence levels and high population frequencies (MAF > 0.05) were excluded.

Lastly, short tandem repeats (STRs) were evaluated using ExpansionHunter (13). Thresholds were defined according to STRipy (https://stripy.org/database)(14) and DRED (http://omicslab.genetics.ac.cn/dred/)(15). Orthogonal confirmation of variants The confirmation of *de novo* variants observed in the trio-ES, especially those that could not be confirmed using IGV, was performed. For this, Sanger sequencing using an Applied Biosystems 3500 Genetic Analyzer (Thermo Fisher Scientific, Foster City, California, USA) was performed. The generated chromatograms were analyzed using the Sequencing Analysis software version 5.4.

## Results

We included 19 unrelated Brazilian individuals with OAFNS (Figure 1), 10 males and 9 females, ranging from 1 to 37 years old. Thirteen of them were reported by Serigatto et al. (2), and six were new cases not previously reported. Mean maternal and paternal age at conception were 27.2 and 32.2 years, respectively. Consanguinity was not observed. Discordant twins were observed in one case. The main clinical features of all 19 cases and respective frequencies were reviewed and updated (Table 1).

**FIgure 1.** Clinical aspects of the individuals with oculoauriculofrontonasal syndrome. A) Case 1; B) Case 4; C) Case 5; D) Case 6; E) Case 14; F) Case 15; G) Case 16; H) Case 17; I) Case 18; J) Case 19.

**Table 1.** Clinical features of the individuals included in the present study.

| **Clinical features** | **1<sup>a</sup>** | **2<sup>a</sup>** | **3<sup>a</sup>** | **4<sup>a</sup>** | **5<sup>a</sup>** | **6<sup>a</sup>** | **7<sup>a</sup>** | **8<sup>a</sup>** | **9<sup>a</sup>** | **10<sup>a</sup>** | **11<sup>a</sup>** | **12<sup>a</sup>** | **13<sup>a</sup>** | **14** | **15** | **16** | **17** | **18** | **19** | **f (%)** |
|---|---|---|---|---|---|---|---|---|---|---|---|---|---|---|---|---|---|---|---|---|
| **Sex** | F | F | M | F | F | M | M | M | F | F | M | M | F | F | M | F | M | M | M | **10M:9F** |
| **Facial asymmetry** | + | + | + | + | + | + | + | + | + | + | + | + | + | + | + | + | + | + | + | **19/19 (100%)** |
| **Ocular hypertelorism** | + | + | + | + | + | + | + | + | + | + | + | + | + | + | + | + | + | + | + | **19/19 (100%)** |
| **Eyelid coloboma** | Up L | - | Up R | Up L | - | Up L/Low R | Up R | - | Up L | Up R | Up L | Up Bi | Up L | - | - | - | - | - | Up L | **11/19 (57.9%)** |
| **Epibulbar dermoid** | + | + | + | - | + | + | - | + | + | + | + | + | + | + | + | + | - | + | + | **16/19 (82.4%)** |
| **Ano/Microphthalmia** | - | - | - | - | - | - | - | - | - | - | - | - | - | - | - | + | - | + | - | **2/19 (10.5%)** |
| **Broad nose** | + | + | + | + | + | + | + | + | + | + | + | + | + | + | + | + | - | - | + | **17/19 (89.4%)** |
| **Bifid/Notched nose** | + | - | - | - | + | + | - | - | + | + | - | - | + | - | - | - | - | - | + | **7/19 (36.8%)** |
| **Nasal polyp** | - | - | + | - | - | - | - | - | - | + | - | + | - | - | + | + | - | - | - | **5/19 (26.3%)** |
| **Cleft lip and palate** | + | CP | + | + | Med | Med | Med | +/Med | + | Med | Med | + | CP/Med | Med | - | + | CP | Med | + | **18/19 (94.7%)** |
| **Rare craniofacial clefts (Tessier)** | 0; 1-13; 2-12; 3-11; 7 | 0; 7 | 0; 2 | 2; 7 | 0-14; 2-12 | 0; 2; 7 | 0 | 0; 2; 7 | 0; 2 | 0; 8 | 0; 2; 10 | 7 | 0-14; 2-12; 6; 7; 8; 10 | 0 | 2 | 7; 8 | 7; 30 | 0 | 1-13; 2; 3; 7; 14 | **19/19 (100%)** |
| **Ear anomalies** | + | + | + | + | - | - | + | + | + | + | + | + | + | - | + | + | + | + | + | **16/19 (82.4%)** |
| **Preauricular tags** | + | + | + | + | + | + | + | + | + | + | + | + | - | + | + | + | + | + | + | **18/19 (94.7%)** |
| **Ectopic nasal bone** | + | - | - | + | + | + | - | - | - | N/E | - | N/E | + | N/E | - | - | - | N/E | + | **6/15 (40.0%)** |
| **CNS anomalies** | + | + | + | - | + | + | - | - | N/E | - | - | N/E | - | N/E | - | - | + | N/E | - | **6/15 (40.0%)** |
| **Costovertebral anomalies** | - | N/E | N/E | - | N/E | + | + | - | + | - | + | + | N/E | N/E |  | - | - | + | + | **7/14 (50.0%)** |
| **Congenital heart defect** | N/E | + | + | N/E | - | N/E | - | + | N/E | N/E | N/E | N/E | + | N/E | - | + | + | + | N/E | **7/10 (70.0%)** |
| **Developmental delay** | - | + | + | - | - | - | - | - | - | - | - | - | - | - | + | - | + | + | - | **5/19 (26.3%)** |
| **Craniofacial involvement** | bilateral | bilateral | bilateral | bilateral | bilateral | bilateral | bilateral | bilateral | bilateral | bilateral | bilateral | bilateral | unilateral | bilateral | bilateral | bilateral | bilateral | unilateral | bilateral | **2 Uni:17 Bi** |
| **Case number in Serigatto et al. (2023)** | 17 | 30 | 29 | 32 | 22 | 25 | 31 | 27 | 2 | 13 | 18 | 28 | 26 | N/R | N/R | N/R | N/R | N/R | N/R |  |

Solo-ES was performed in 16 cases and trio-ES in 3. One of our previously published cases (Case 13) had been sequenced and showed negative results(5); however, the data were updated and annotated to allow a comparison with the ES data obtained from all the remaining cases.

For the analysis of all probands, including those that also had available parental samples (trio-ES), firstly, we aimed to exclude blended phenotypes or overlapping conditions with known molecular basis by analyzing potential causal variants associated with OMIM phenotypes (Supplementary Table 1). Secondly, we searched for genes presenting very rare variants in common to two or more individuals, with and without associated phenotypes (Supplementary Table 2). No variant or candidate gene was found among our samples. The same variant was observed in two or more individuals in a few instances but was not associated with the phenotype.

For the trio-ES analysis, the first step included the same analysis performed in the solo- ES analysis. Later, the analysis focused on *de novo* variants present in the affected proband. Of note, none of the parents had OAFNS; however, in one family in which trio-ES was done, an upper incisor diastema and a mild form of midline cleft affecting the alveolus (confirmed by a periapical X-ray) were observed in the father, without other anomalies. Trio-ES analysis revealed *de novo* variants absent in population databases, but none of them could be associated with the phenotype of OAFNS. In addition, these *de novo* variants were exclusively observed in each family analyzed and, therefore, not considered as candidates. Furthermore, none of the *de novo* variants identified in trio-ES were found in the remaining cases that underwent solo-ES.

The CNVs and STRs analysis were unremarkable. Case 5 had an abnormal karyotype (47,XX, +mar); however, the CNV analysis through ES could not identify the origin of this marker chromosome.

Lastly, the secondary findings, according to the updated ACMG guidelines (v3.3), were not found to be reportable in any case.

## Discussion

In this case series, we included 19 individuals with OAFNS, including six unpublished cases. Clinical data of 13 individuals (Cases 1-13) had already been reported elsewhere (2,5); nonetheless, molecular investigations were not performed at that time, except for Case 13, which had already undergone ES (5).

Briefly, the clinical findings of the six new cases (Cases 14-19), four males and two females, are in accordance with the literature (1,2). Five of them had bilateral involvement of the craniofacial structures, whereas only one had unilateral involvement. The exclusive finding of OAFNS, namely the ectopic nasal bone, was observed in only one of the six new cases investigated through skull CT scans, present as a bone bridge connecting the maxilla to the nasal bones, similar to other reports from the literature (5,7).

Currently, genetic investigations performed in individuals with OAFNS are limited to a few sporadic cases. Segregation analysis stated that an autosomal recessive pattern is unlikely but did not exclude the possibility of an autosomal dominant inheritance (16), although no recurrence has been reported in the literature. Karyotyping was done in approximately 50% of the cases and showed abnormalities in only one case (2,7). Chromosomal microarray analysis was negative in four cases (2,17,18). Sanger sequencing of ALX-related FND genes (*ALX1, ALX3* and *ALX4*) performed in previous studies showed no variants (1). Prior to this study, trio-ES had been performed in four individuals with OAFNS and did not find variants associated with the phenotype (1). One of our cases (Case 13) has already undergone solo-ES, but it was negative (5). Furthermore, individuals presenting overlapping features of Pai syndrome (OMIM 155145) and OAFNS were screened by ES, and no causal variants were identified, except for a maternally inherited *GLI2* variant, classified as VUS, that was reported considering the hypothesis of incomplete penetrance (6). Therefore, cytogenetic and molecular investigations have not been capable of elucidating the etiology of OAFNS yet, including the present study, since no pathogenic/likely pathogenic variants were identified and no candidate genes were suggested through the ES of 19 individuals with OAFNS.

Our analysis comparing genetic findings from the 19 probands, focusing on similarities, either by finding the exact same rare variant in two or more cases or by searching different rare variants within the same gene in two or more cases, was not elucidating (Supplementary Table 2). In the former situation, the same variant was only observed in multiple cases when its population frequency was high and, therefore, characterized a common variant. The latter scenario resulted in few genes that contained two or more variants in different cases, but none of them could be associated with the phenotype, owing to their inconsistency with the inheritance model, a very distinct associated OMIM phenotype, a distinct signaling pathway unrelated to craniofacial development, or even the gene function, making it unfeasible to suggest candidate genes based on the available data.

Focusing on *de novo* variants, our trio-ES analysis retrieved three heterozygous variants of interest, one in each family. The variant NM_003565.4:c.2174T>G in *ULK1* was absent in both parents and had a CADD score of 28.1. The *ULK1* (Unc-51 Like Autophagy Activating Kinase 1; OMIM 603168) gene encodes a serine/threonine-protein kinase that initiates autophagosome formation and induces autophagy in response to starvation. Also, ULK1 inhibits the kinase activity of mTORC1 and cell proliferation. We could not confirm this variant by Sanger sequencing. Nevertheless, the role of this variant is uncertain. The variant NM_006364.4:c.1901C>T in *SEC23A* was absent in both parents, was very rare in population databases, was observed at a 20% VAF and confirmed to be a mosaic by Sanger sequencing, predicted as deleterious, and had a CADD score of 33. This gene encodes SEC23 homolog A (OMIM 610511), an essential component of coat protein complex II that functions in vesicle budding from the endoplasmic reticulum and endoplasmic reticulum-to-Golgi protein trafficking. Pathogenic variants in this gene are associated with craniolenticulosutural dysplasia (OMIM 607812), congenital disorder of glycosylation type In, and amelogenesis imperfecta type Ie, and have been linked to bone disease. Although there are few overlapping features with craniolenticulosutural dysplasia, this variant does not explain the full picture of OAFNS because there is no involvement of the same developmental fields, e.g., frontonasal process and first and second pharyngeal arches. The variant NM_024308.4:c.415C>T in *DHRS11* was absent in both parents and was confirmed by Sanger sequencing. This variant is very rare in population databases, was predicted as deleterious, and had a CADD score of 27.7. DHRS11 (Dehydrogenase/Reductase 11; OMIM 616159) encodes a short-chain dehydrogenase/reductase that catalyzes steroid and xenobiotic ketone reduction and contributes to androgen activation. Variants in *DHRS11* are associated with steroid hormone-related and metabolic disease themes. Therefore, no association with the phenotype of OAFNS was observed, owing to its function and expression. Regarding the family in which the father had an alveolar midline cleft, we also analyzed variants that were common to the proband, considering the hypothesis of a probable incomplete penetrance, but none of them provided enough evidence to assume any association. Thus, this finding could be considered “fortuitous”.

Considering the significant overlap of OAFNS with FND and CFM, we also analyzed known variants that could cause both phenotypes independently and result in the full picture of OAFNS, but no variants were found. FND is a diverse and heterogeneous group, encompassing several rare conditions, and its etiology is unclear in most cases (19,20). It can also occur as an isolated field defect or associated with other anomalies (21). The aristaless-like homeobox family has three members that are associated with FND, all consistent with autosomal recessive inheritance: *ALX3* is associated with FND type 1 (OMIM 136760), also known as frontorhiny, *ALX4* is associated with FND type 2 (OMIM 605420), and *ALX1* is associated with FND type 3 (OMIM 613456). Screening of these three *ALX* genes in the present study did not identify causative variants. Other examples of conditions associated with FND, and known molecular basis, include the acromelic frontonasal dysostosis (OMIM 603671) caused by heterozygous variants in *ZSWIM6*(22), the acrofrontofacionasal dysostosis (OMIM 201180; 237910) caused by homozygous variants in *PIGB*(23), the craniofrontonasal syndrome (OMIM 304110) caused by heterozygous variants in *EFNB1*(24,25). These conditions share features that characterize FND, but each presents other unique phenotypic features that allow them to be clinically distinguished and, therefore, have different genetic causes, as well as different pathways and mechanisms involved in their pathogenesis (19,20). Furthermore, there are well-defined conditions presenting FND as a feature that have clinically recognizable patterns but unknown etiology. An interesting example, as it is the main differential diagnosis of OAFNS, is the Pai syndrome, a rare condition characterized by ocular hypertelorism, median cleft lip, nasal and/or facial polyps, and pericallosal lipoma. Research conducted by independent groups could not elucidate the etiology of Pai syndrome (6,26) through NGS and also by sequencing affected tissue, namely nasal polyp, to test the hypothesis of mosaicism, and negative results led to this condition being included in the list of unsolvable rare diseases (6). Together, these data illustrate the heterogeneous genetic landscape of conditions characterized by FND and underscore that clinically well-defined conditions still have obscure etiologies.

In relation to CFM, also known as the oculoauriculovertebral spectrum, it is a clinically and etiologically heterogeneous condition characterized by mandibular hypoplasia, ear anomalies, and preauricular tags. Currently, there are two principal genes associated with CFM: *SF3B2* (Splicing factor 3B, subunit 2) and *FOXI3* (Forkhead box I3)(27–29). Variants in *SF3B2* leading to haploinsufficiency represent the most prevalent genetic cause of CFM in sporadic and familial cases. *SF3B2* knockdown in *Xenopus laevis* disrupts the formation of cranial neural crest precursors and leads to craniofacial cartilage abnormalities, reinforcing the association between spliceosome defects and abnormal neural crest development in congenital craniofacial disorders (27). Remarkably, spliceosomal defects have been associated with several rare craniofacial disorders, a group termed craniofacial spliceosomopathies, including Nager syndrome (OMIM 154400), mandibulofacial dysostosis Guion-Almeida type (OMIM 610536), cerebrocostomandibular syndrome (OMIM 117650), Burn-McKeown syndrome (OMIM 608572), and Richieri-Costa-Pereira syndrome (OMIM 268305), that have common features such as mandibular hypoplasia and ear anomalies (30,31). The other CFM-associated gene, *FOXI3,* is expressed in the preplacode region and in the endoderm and ectoderm of the pharyngeal arches, playing an important role in the formation of the inner ear and in craniofacial development, especially of the mandible (29,32). In addition, Foxi3 is involved in the FGF, Wnt, and BMP signaling pathways, pathways that play a role in the differentiation of the pharyngeal arches (32). Moreover, other genes less frequently associated with CFM were also investigated (33).

Interestingly, both FND and CFM were included in the group of neurocristopathies (NCP), a diverse group of disorders that arise from abnormalities in the neural crest cells (NCCs) during embryonic development. The NCCs are a transient, multipotent, and migratory embryonic population, often termed the fourth germinative layer, that emerges at the boundary of the neural and non-neural ectoderm, representing a diverse array of specialized cells and tissues across the entire body, involved in distinct developmental fields. Their formation results from a complex, multistep, and time-regulated process that is coordinated by a network of signaling pathways including Wnt, BMP, FGF, Hedgehog, and retinoic acid (34,35). Notably, cranial NCCs are responsible for the origin of the frontonasal process and the pharyngeal arches. Therefore, FND and CFM have already been suggested to represent examples of NCP, even before the elucidation of their etiology. CFM has been considered a NCP owing to its abnormalities arising mainly from the first and second pharyngeal arches, along with ocular and skeletal anomalies, all of which are derivatives of the NCCs. The same fits FND, whose anomalies arise from abnormal development of the frontonasal process, which originates from the cranial NCCs. Furthermore, NP can be classified into three types, considering the stage of development in which the disturbance of NCCs occurs: (i) induction and specification, (ii) migration, and (iii) differentiation (36). Accordingly, CFM would be classified as a NCP resulting from induction and specification, as observed in animal models (27), but also as a migration and differentiation defect, since the abnormally developed NCCs still populate the pharyngeal arches and undergo cellular differentiation, resulting in abnormalities in their derivatives. Since the widespread exposure window observed from the formation of NCCs until the development of the pharyngeal arches, it may be susceptible to various factors at different stages of development that could explain the heterogeneity and clinical variability of CFM. The same can be said about FND, which seems to be a NCP resulting from a migration defect, where the NCCs are normally developed; however, they fail to migrate to their destination (19,37). These observations may pave the way for future approaches regarding the etiology of OAFNS, which may be caused by early disturbance in the development of the cranial NCCs and their derivatives.

Facing previous negative results (1,5,6), and adding further evidence provided here, the etiology of OAFNS tends towards non-traditional inheritance models. After increasing the number of individuals with OAFNS evaluated by NGS and providing sufficient data for analysis, we can exclude monogenic inheritance and support alternative hypotheses that have been raised before, such as oligogenism, tissue mosaicism, methylation defects, and non-genetic causes (1). Therefore, multi-omics approaches should be included in further studies. These approaches could provide evidence of possible early disturbance in the development of NCCs by analyzing signaling pathways and genes involved in this process, beyond the ES data. With the current advances in long-read GS, trio analysis using this technique could be elucidating in this scenario, either by covering the whole genome with accuracy or by analyzing methylation profiles.

Altogether, the analysis of 19 individuals with OAFNS in the present study allows us to exclude coding variants in exonic regions that follow Mendelian inheritance as possible causes of OAFNS. Heterozygous, *de novo* variants are also unlikely to be causative, at least in an autosomal dominant inheritance, yet not excluding the hypothesis of oligogenism. Limitations of our analysis include variants present in regions not covered by ES, and the fact that trio-ES analysis was not done in all cases due to the lack of parental samples. Further approaches should include long-read GS to uncover regions not evaluated by ES and analyze methylation profiles through the same technique, and other omics, such as transcriptomics and proteomics, to investigate non-traditional inheritance models. Also, analysis of affected tissue should be performed to test the hypothesis of mosaicism, which has not been fully ruled out.

## Data Availability Statement

The authors declare that the data supporting the findings of this study are available within the paper and its Supplementary Information files. Should any raw data files be needed in another format they are available from the corresponding author upon reasonable request.

## Acknowledgments

The authors acknowledge the São Paulo Research Foundation (FAPESP), Brazil – Process Number 2023/15095-9 and 2024/02970-1 – and the Coordenação de Aperfeiçoamento de Pessoal de Nível Superior - Brazil (CAPES).

## Author contribution statement

H. R. Serigatto: conceptualization, methodology, analysis, investigation, resources, writing – original draft, writing – review and editing, visualization, funding acquisition. N. M. Kokitsu-Nakata: investigation, resources. S. Vendramini-Pittoli: investigation, resources. C. Tonello: investigation, resources. M. R. Passos-Bueno: methodology, resources. R. M. Zechi-Ceide: conceptualization, methodology, investigation, resources, writing – review and editing, supervision, project administration, funding acquisition.

